# Predictors of mortality in hospitalized COVID-19 patients in Athens, Greece

**DOI:** 10.1101/2020.10.12.20211193

**Authors:** Dimitrios Giannoglou, Evangelia Meimeti, Xenia Provatopoulou, Konstantinos Stathopoulos, Ioannis –Kriton Roukas, Petros Galanis

## Abstract

**Background:** The epidemic of COVID-19 has rapidly spread worldwide, with millions of confirmed cases and related deaths. Numerous efforts are being made to clarify how the infection progresses and potential factors associated with disease severity and mortality. We investigated the mortality in Greek hospitalized COVID-19 patients and also the predictors of this mortality.

**Methods:** Study population included 512 COVID-19 patients admitted to the hospitals of the Attica region of Greece. Patients’ demographic characteristics, comorbidities, allergies, previous vaccination for seasonal influenza virus, admission to ICU, intubation, and death were recorded. Potential predictors of in-hospital mortality were identified by regression analysis.

**Results:** The mean age of hospitalized patients was 60.4 years, and was higher in patients who deceased. The most common comorbidities were respiratory diseases, hypertension, gastrointestinal disorders, dyslipidemia, mental health diseases, asthma, diabetes mellitus and cardiovascular diseases. The need for ICU care and intubation was significantly higher among patients who died. The mortality rate was 15.8% (81 out of 512). Age ≥65 years, cancer, chronic kidney disease, endocrine diseases, central nervous system disorders, anemia, and intubation were independently associated with increased in-hospital mortality, while allergies and previous influenza vaccination were associated with decreased in-hospital mortality.

**Conclusion:** Our finding of a beneficial effect of allergies and influenza vaccination against COVID-19 infection merits further investigation, as it may shed light in the mechanisms underlying disease progression and severity. Most importantly, it may assist in the implementation of efficient protective measures and public healthcare policies.

## Introduction

The epidemic of coronavirus disease 2019 (COVID-19) originally emerged in Wuhan, China in late December 2019, and rapidly spread worldwide. By the end of June 2020, the global number of confirmed cases had reached 9,952,507, and COVID-19-related deaths were 498,519 (1). In European countries, 2,593,558 confirmed cases and 195,889 deaths had been reported during the same time period, with Europe ranking second following America. Significant variations both in confirmed cases and in mortality rates are observed between European countries (1). Whether these reflect the extent and effect of the applied public health control measures, the availability and access to healthcare facilities, the overall health state or a potential genetic susceptibility background of each population, the curve of infection or other potential contributing factors is worthy of further investigation.

Although COVID-19 infection is typically of mild symptomatology with most patients fully recovering, critically ill patients require long-term hospitalization and are in a considerable risk of death, mainly due to progressive respiratory failure from lung infection of severe acute respiratory syndrome coronavirus 2 (SARSCoV-2) (2). Data so far suggest that SARS-CoV-2 might directly dysregulate kidney and liver normal function as well as peripheral blood components, ultimately leading to multiple organ failure and death (2-4). Mortality from COVID-19 has been systematically associated with older age, and the presence of comorbidities, primarily hypertension, diabetes, obesity, and cardiovascular and lung disease in many different populations (5). To our knowledge, this is the first study in Greece that investigates the mortality in COVID-19 patients in Greece and also the predictors of this mortality.

## Methods

The administrative region of Attica is the largest region of Greece, encompassing the entire metropolitan area of the capital city of Athens as well as nearby cities. Its population is approximately 3.83 million people, accounting for 35.4% of the total country population (6). During the period 21/2/2020 and 30/6/2020, a total of 512 patients diagnosed with emergent COVID-19 infection were admitted to 14 General Hospitals pertaining to the 1st Regional Health Authority of Attica. For all patients, data were recorded including their demographic characteristics, comorbidities, previous vaccination for seasonal influenza virus, need for admission to ICU and for intubation, and patient outcome (discharge or death).

The study was conducted in accordance to the Helsinki Declaration for the ethical principles for medical research involving human subjects. Data were provided by the 1st Regional Health Authority of Attica, following the approval of the protocol by the Scientific Committee of the Greek Ministry of Health (27628 / 23-06-2020), ensuring legality of conduct, compliance with medical ethical standards and scientific validity.

### Statistical analysis

Continuous variables are presented as mean (standard deviation) and categorical variables are presented as numbers (percentages). Age was divided in 10-years intervals in order to calculate mortality by these intervals and gender in hospitalized COVID-19 patients. We included gender, age, influenza vaccination, ICU care, intubation and comorbidities in regression models. First, we performed univariate logistic regression analysis and then variables that were significantly different (p<0.20) were entered into the backward stepwise multivariate logistic regression analysis. Multivariate logistic regression analysis was applied for the control of each potentially confounding of each statistically significant predictive factor to the others. We estimated adjusted odds ratios (OR) with 95% confidence intervals (CI) and p-values. All tests of statistical significance were two-tailed, and p-values<0.05 were considered significant. Statistical analysis was performed with the Statistical Package for Social Sciences software (IBM Corp. Released 2012. IBM SPSS Statistics for Windows, Version 21.0. Armonk, NY: IBM Corp.).

## Results

Study population included 512 hospitalized COVID-19 patients and demographic characteristics and comorbidities of patients are shown in Table 1. Mean age was 60.4 years (standard deviation, 18.2) and was higher in patients who died than patients discharged alive (75.8 vs. 57.5 years). Twenty-five point four percent of the patients were vaccinated for influenza. The most common comorbidities were respiratory diseases (38.3%), hypertension (33.6%), gastrointestinal disorders (24.4%), dyslipidemia (22.5%), mental health diseases (20.1%), asthma (13.9%), diabetes mellitus (11.9%), coronary artery disease (11.7%) and heart arrhythmias (10.4%). The need for ICU care and intubation was 18.9% and 14.8 respectively. Of those patients who died, almost half were hospitalized in ICU (56.8%) and were intubated (49.4%).

**Table 1.** Demographic characteristics and comorbidities in hospitalized COVID-19 patients.

| **Demographic characteristics** | **Discharged, N (%) =**<br>**431 (84.2%)<sup>a</sup>** | **Died, N (%) =**<br>**81 (15.8%)<sup>b</sup>** | **Total population, N (%) =**<br>**512 (100%)<sup>c</sup>** |
| --- | --- | --- | --- |
| Females | 170 (39.4) | 25 (30.9) | 195 (38.1) |
| Age (mean, standard deviation) (years) | 57.5 (17.5) | 75.8 (13.1) | 60.4 (18.2) |
| Influenza vaccine | 103 (23.9) | 27 (33.3) | 130 (25.4) |
| **Comorbidities** |  |  |  |
| Coronary artery disease | 37 (8.6) | 23 (28.4) | 60 (11.7) |
| Congestive heart failure | 30 (7.0) | 13 (16.0) | 43 (8.4) |
| Hypertension | 124 (28.8) | 48 (59.3) | 172 (33.6) |
| Heart arrhythmias | 39 (9.0) | 14 (17.3) | 53 (10.4) |
| Dyslipidemia | 95 (22.0) | 20 (24.7) | 115 (22.5) |
| Peripheral vascular disease | 35 (8.1) | 5 (6.2) | 40 (7.8) |
| Diabetes mellitus | 45 (10.4) | 16 (19.8) | 61 (11.9) |
| Cancer | 22 (5.1) | 13 (16.0) | 35 (6.8) |
| Respiratory diseases (bronchitis, pneumonia, infections) | 170 (39.4) | 26 (32.1) | 196 (38.3) |
| Asthma | 61 (14.2) | 10 (12.3) | 71 (13.9) |
| Allergies | 32 (7.4) | 4 (4.9) | 36 (7.0) |
| Chronic kidney disease | 13 (3.0) | 13 (16.0) | 26 (5.1) |
| Endocrine diseases | 25 (5.8) | 15 (18.5) | 40 (7.8) |
| Disorders of purine and pyrimidine metabolism | 12 (2.8) | 9 (11.1) | 21 (4.1) |
| Central nervous system disorders | 20 (4.6) | 11 (13.6) | 31 (6.1) |
| Anemia | 20 (4.6) | 18 (22.2) | 38 (7.4) |
| Gastrointestinal disorders | 92 (21.3) | 33 (40.7) | 125 (24.4) |
| Mental health diseases | 85 (19.7) | 18 (22.2) | 103 (20.1) |
| ICU care | 51 (11.8) | 46 (56.8) | 97 (18.9) |
| Intubation | 36 (8.4) | 40 (49.4) | 76 (14.8) |

<sup>a</sup> Percentage based on denominator N =512, remaining percentages in this column are based on denominator N = 431
<sup>b</sup> Percentage based on denominator N =512, remaining percentages in this column are based on denominator N = 81
<sup>c</sup> Percentages in this column are based on denominator N = 512

Out of the 512 patients, 81 died (15.8%) and 431 were discharged alive (84.2%). Mortality by gender and 10-years age interval in patients is shown in Table 2. Males had higher mortality than females (17.7% vs. 12.8%). Also, increased age was associated with increased mortality. In particular, only one patient <40 years of age died, while the mortality in <50 years was 2.3% and in ≥50 years was 20.4%. Mortality was highest in the 80-89-years age group (42.6%) and in >89 years (42.3%).

**Table 2.** Mortality by gender and 10-years age intervals in hospitalized COVID-19 patients.

| Age interval (years) | Discharged, N (%) |  | Died, N (%) |  | Mortality, % |
| --- | --- | --- | --- | --- | --- |
|  | Males | Females | Males | Females |  |
| <20 | 6 (100) | NA | 0 (0) | NA | 0 |
| 20-29 | 19 (100) | 10 (100.0) | 0 (0) | 0 (0) | 0 |
| 30-39 | 14 (93.3) | 16 (100.0) | 1 (6.7) | 0 (0) | 3.2 |
| 40-49 | 42 (95.5) | 22 (100.0) | 2 (4.5) | 0 (0) | 3.0 |
| 50-59 | 59 (93.7) | 42 (95.5) | 4 (6.3) | 2 (4.5) | 5.6 |
| 60-69 | 58 (84.1) | 28 (90.3) | 11 (15.9) | 3 (9.7) | 14.0 |
| 70-79 | 40 (71.4) | 28 (80.0) | 16 (28.6) | 7 (20.0) | 25.3 |
| 80-89 | 18 (52.9) | 13 (65.0) | 16 (47.1) | 7 (35.0) | 42.6 |
| >89 | 4 (40.0) | 11 (68.8) | 6 (60.0) | 5 (31.3) | 42.3 |

NA: non applicable

Univariate and multivariate logistic regression analyses for in-hospital mortality are shown in Table 3. According to multivariate analysis, age ≥65 (OR = 8.22, 95% CI = 3.88-17.42), cancer (OR = 3.76, 95% CI = 1.39-10.20), chronic kidney disease (OR = 5.17, 95% CI = 1.41-18.91), endocrine diseases (OR = 3.13, 95% CI = 1.19-8.27), central nervous system disorders (OR = 5.73, 95% CI = 2.13-15.43), anemia (OR = 8.32, 95% CI = 3.1-22.35), and intubation (OR = 8.08, 95% CI = 1.89-34.6) were independently associated with increased in-hospital mortality. On the contrary, influenza vaccine (OR = 0.38, 95% CI = 0.17-0.81) and allergies (OR = 0.16, 95% CI = 0.04-0.67) were independently associated with decreased in-hospital mortality.

**Table 3.** Univariate and multivariate logistic regression analyses for increasing risk of in-hospital mortality.

| Variable | Unadjusted OR (95% CI) | P-value | Adjusted OR (95% CI)<sup>a</sup> | P-value |
| --- | --- | --- | --- | --- |
| Males vs. females | 1.46 (0.88 – 2.43) | 0.15 | NS |  |
| Age ≥65 vs. age <65 years | 6.85 (3.87 – 12.12) | <0.001 | 8.22 (3.88 – 17.42) | <0.001 |
| Influenza vaccine | 1.59 (0.95 – 2.66) | 0.075 | 0.38 (0.17 – 0.81) | 0.013 |
| Coronary artery disease | 4.22 (2.34 – 7.61) | <0.001 | NS |  |
| Congestive heart failure | 2.56 (1.27 – 5.15) | 0.009 | NS |  |
| Hypertension | 3.60 (2.21 – 5.88) | <0.001 | NS |  |
| Heart arrhythmias | 2.10 (1.08 – 4.08) | 0.028 | NS |  |
| Dyslipidemia | 1.16 (0.67 – 2.02) | 0.60 | NS |  |
| Peripheral vascular disease | 0.74 (0.28 – 1.96) | 0.55 | NS |  |
| Diabetes mellitus | 2.11 (1.13 – 3.96) | 0.02 | NS |  |
| Cancer | 3.55 (1.71 – 7.39) | 0.001 | 3.76 (1.39 – 10.20) | 0.009 |
| Respiratory diseases | 0.73 (0.44 – 1.20) | 0.21 | NS |  |
| Asthma | 0.85 (0.42 – 1.75) | 0.67 | NS |  |
| Allergies | 0.65 (0.22 – 1.89) | 0.43 | 0.16 (0.04 – 0.67) | 0.011 |
| Chronic kidney disease | 6.15 (2.73 – 13.82) | <0.001 | 5.17 (1.41 – 18.91) | 0.013 |
| Endocrine diseases | 3.69 (1.85 – 7.37) | <0.001 | 3.13 (1.19 – 8.27) | 0.021 |
| Disorders of purine and pyrimidine metabolism | 4.37 (1.78 – 10.73) | 0.001 | NS |  |
| Central nervous system disorders | 3.23 (1.48 – 7.03) | 0.003 | 5.73 (2.13 – 15.43) | 0.001 |
| Anemia | 5.87 (2.95 – 11.70) | <0.001 | 8.32 (3.10 – 22.35) | <0.001 |
| Gastrointestinal disorders | 2.53 (1.54 – 4.18) | <0.001 | NS |  |
| Mental health diseases | 1.16 (0.65 – 2.07) | 0.61 | NS |  |
| ICU care | 9.79 (5.78 – 16.60) | <0.001 | NS |  |
| Intubation | 10.71 (6.16 – 18.62) | <0.001 | 8.08 (1.89 – 34.60) | 0.005 |

CI: confidence interval
NS: not selected by the backward elimination procedure in the multivariate logistic regression analysis with a significance level set at 0.05
<sup>a</sup> R<sup>2</sup> for the final multivariate model was 50%

## Discussion

In our study, ICU admission and intubation were applied to 18.9% and 14.8% of patients, respectively, indicating increased disease severity. As expected, both ICU admission and intubation were far more common among patients who died compared to those discharged (56.8% vs. 11.8%, and 49.4% vs. 8.4%, respectively). A previous study in 102 patients hospitalized with COVID-19 infection to Wuhan University Zhongnan Hospital indicates a similar percentage of patients (17.6%) admitted to the ICU. In agreement with our observations, ICU admission was less frequent in hospitalized patients who were discharged (14.1% vs. 35.3%) (7).

It comes as no surprise that among hospitalized patients, the vast majority were males, of older age and suffering from various comorbidities, primarily respiratory diseases hypertension, gastrointestinal disorders, dyslipidemia, mental diseases, asthma, diabetes mellitus and cardiovascular diseases. An overall analysis of 1,458 COVID-19 patients from 5 studies showed that the most common comorbidities are hypertension (55.3%), cardiovascular and cerebrovascular disease (31.5%), and diabetes (30.6%), followed by hepatitis and liver cirrhosis, malignancy, chronic obstructive pulmonary disease, chronic renal failure, gastrointestinal disorders, immunodeficiency and diseases of the central nervous system at much lower rates (8).

We found that increased age was associated with increased mortality. Notably, mortality was exceptionally high in patients ≥80 years of age (42.6%) whereas no fatality was observed in patients <29 years of age. In agreement, a study in 4226 patients with COVID-19 in the United States also indicated highest mortality rates in patients older than 85 years, followed by patients aged 55 to 84 years, while zero mortality was reported among patients 19 years and younger (9). The close association between increasing age, particularly >80 years, and mortality from COVID-19 infection has also been verified in a large series of 44,672 confirmed cases from China (10).

Multivariate analysis identified that comorbidities i.e. cancer, chronic kidney disease, endocrine disease, central nervous system disease and anemia were associated with increased mortality. Preliminary reports from China suggested very early that cardiovascular risk factors such as hypertension, diabetes, obesity and established cardiovascular disease were common amongst hospitalized patients with COVID-19, and were associated with a high risk of in-hospital mortality (3, 11-13). These observations were verified in patients in Northern Italy, with mortality rates of 36% in hospitalized COVID-19 patients with cardiac disease compared to 15% to those without (14). Another study from Wulan also showed than among hospitalized patients, those who survived were younger and were less likely to suffer from hypertension, diabetes and chronic kidney disease (8). In a series of 191 patients with laboratory-confirmed COVID-19, mortality increased with age, sequential organ failure assessment and comorbidities including hypertension, diabetes and coronary heart disease (15). Similarly, data from 239 critically ill COVID-19 patients from 3 hospitals in China revealed that age >65 years, acute respiratory distress syndrome, acute cardiac injury, acute kidney injury, liver dysfunction and coagulopathy were associated with increased mortality (16). A study with 8,910 COVID-19 hospitalized patients in 169 hospitals in Asia, Europe, and North America found that age >65 years, coronary artery disease, congestive heart failure, cardiac arrhythmia, chronic obstructive pulmonary disease and smoking were independent factors of mortality (17). Cardiovascular and respiratory diseases did not come up as potential risk factors of mortality from COVID-19 in our study population. This could potentially be attributed to the relatively small sample size in combination with the high prevalence of these diseases in the general Greek population, particularly of older age. Notably, cardiovascular events and respiratory diseases rise as the first and fourth cause of mortality in the general population, respectively (18). Regarding cancer and COVID-19 infection, data from a cohort study on patients from the USA, Canada, and Spain from the COVID-19 and Cancer Consortium (CCC19) database indicated an increased 30-day all-cause mortality, associated with general risk factors and risk factors unique to cancer patients (increased age, male sex, smoking, cancer status, ECOG performance status and the presence of two or more comorbidities requiring treatment) (19). Patients with cancer not only had a higher risk of developing more serious infection but they also had a more accelerated health deterioration (20). In support, research in a tertiary hospital in Wuhan found that 25% of patients with cancer and SARS-COV-2 infection died, most of them over 60 years of age (21). As far as the presence of endocrine diseases, diseases of the central nervous system and anemia, they are not typically included among the potential risk factors investigated in most studies. Thus, differences between studies in data collection and reporting of comorbidities does not allow for reliable comparisons to be made.

In our study, intubation came up as a risk factor in our multivariate model. An increased need for treatment in ICU has been reported for patients with COVID-19 infection and pre-existing cardiac disease (14). Also, 14% of patients with pre-existing cancer were admitted to the ICU, while 12% required mechanical ventilation (19). Another study in 138 hospitalized patients with COVID-19 indicated that 26.1% of patients were transferred to the ICU owing to complications, including acute respiratory distress syndrome, arrhythmias and shock (31%). Notably, 46.4% of patients had one or more comorbidities including hypertension (31%), diabetes (10%), cardiovascular disease (14.5%), and malignancy (7.2%) (11). According to a comprehensive review, acute respiratory distress syndrome which may further lead to septic shock are the two main complications contributing to ICU care and mortality from COVID-19 in patients older than 60 years, with a history of smoking and comorbid conditions (7). It has been proposed that smoking and older age are associated with a higher density of ACE2 receptors, partially explaining these observations. Another study found that 10-32% of hospitalized patients required admission to the ICU due to respiratory deterioration (22). As far as intubation is concerned, data from China indicate that it was required in approximately 3.2% of patients with COVID-19 (23). According to data from the US, this percentage rises to 18-33% among hospitalized patients, of whom up to 20% die (24-27). Interestingly, a recent analysis revealed a close association between SARS-CoV-2 viral load at admission in hospitalized patients with COVID-19 and the risk of intubation and in-hospital mortality (28).

Interestingly, our data indicate that pre-existing allergies as well as previous vaccination for influenza were independently associated with decreased in-hospital mortality from COVID-19. Allergy is an immune response to antigen stimulation, inducing the release and production of various inflammatory mediators at different organs, however its evolvement in COVID-19 severity and progression is currently unknown. One compiled data analysis in 65 patients with laboratory-confirmed COVID-19 and allergies indicated a more moderate severity of disease (29). A trend towards increased counts of lymphocytes and monocytes as well as higher levels of CD3+, CD4+, and CD8+ T cells were observed in these patients. As COVID-19 severity has been correlated with a significant decrease in lymphocytes (3, 12, 30), these observations are consistent with the finding of a more moderate disease. It was proposed that resistance to SARS-CoV-2 by high-sensitivity state may result in a lighter attack on lymphocytes, inducing the severity of the initial condition of COVID-19 patients (29). Previous studies have suggested that although T cells, B cells and NK cells might all be involved in the immune response to infection by COVID-19, T cells may exert a more important role, with the degree of their reduction being predictive of the progression of COVID-19 (2, 31). Based on existing findings, it has been suggested that the reduction in lymphocyte count in COVID-19 patients with allergies may primarily affect T cells, rather than B or NK cells (29). Although there are some evidence that existing allergies may reduce the destructive power of SARS-CoV-2 infection, further research is necessary to clarify their potential effect and elucidate the underlying mechanisms.

It is widely recognized that influenza vaccination is associated with a substantial reduction in mortality in the general population, notably among the elderly, even after adjustment for typical confounders such as age, sex and comorbidity status (32). According to a recent analysis, an association between COVID-19 severity and influenza vaccination, which appeared protective, was found (33). This beneficial effect was also supported by case incidence and recovery parameters. Similarly, a beneficial relationship between lower respiratory tract infections and COVID-19 severity was observed in countries where influenza immunization is less common (33). The reported data are in support of our observation of a protective effect of previous influenza vaccination from mortality from COVID-19 in our patients. Indeed, a beneficial effect of influenza vaccination on mortality from COVID-19 was recently reported in the elderly US population (34). More specifically, higher influenza vaccination coverage was associated with lower COVID-19 mortality rates, with an average 28% decrease in mortality for every 10% increase in influenza vaccination coverage. Moreover the protective effect appeared to be non-linear, becoming stronger with higher vaccination coverage. These data are further supported by the analysis of 92,664 clinically and molecularly confirmed COVID-19 cases in Brazil (35). According to the results, patients who had been recently vaccinated for influenza experienced 8% lower odds of needing intensive care treatment, 18% lower odds of requiring invasive respiratory support and 17% lower odds of death. Mortality was consistently lower among influenza vaccinated patients across all age groups, with statistically significant difference reported over the age of 30 years. An interesting observation was that the protective effect was evident not only in patients vaccinated prior to COVID-19 infection, but also in those vaccinated after the onset of clinical COVID-19 symptoms, with 20% and 27% lower odds of mortality respectively (35). Altogether these findings suggest that influenza vaccination may potentially play a protective role in COVID-19, with vaccinated patients having higher chances of survival and less need for intensive hospital care. Nevertheless additional confirmatory studies are urgently required to verify these indications.

Although the mechanisms potentially underlining the protective effect of influenza vaccination against COVID-19 infection extend beyond the scope of this work, it has been proposed that the protection may be driven by innate immune changes triggered by natural infections and vaccines with off-target effect against a variety of pathogens (35, 36). Given that SARS-CoV-2 and influenza viruses are evolutionary close, and share similarities with respect to viral structure, epitopes, transmission and pathogenic mechanisms, they may be detected by similar or identical pattern recognition receptors, triggering inflammatory and antiviral responses that involve T cell and NK cell activation and early respiratory IL12p40 and IFN-I responses (33, 37, 38). Along the same lines, recent research has demonstrated that common human coronaviruses like OC43, HKU1, 229E, and NL63, are capable of inducing immune memory against SARS-CoV-2 through CD8+ T cells by sharing antigen epitopes for presentation to the immune system by MHC class I, proposing a potential vaccination strategy for non-specific stimulation of the immune system against COVID-19 (39). Another potential theory proposed to explain the putative protective effect of influenza vaccination against COVID-19 relies on the fact that unvaccinated individuals are at risk of persistent viral infections that lead to a decline in T cell diversity impairing the immune response against other pathogens. Unlike natural infection, influenza vaccination does not induce a strong, virus-specific CD8+ T cell immune response, and this may assist in more efficient clearance of SARS-CoV-2 infection as vaccinated individuals will have more T cell diversity compared with those with natural influenza infection (34).

Our study bears limitations, particularly with regard to the relative small sample size. Nevertheless, we considered the total population of the Athens metropolitan area that required hospitalization for infection by COVID-19 during the period 21/2/2020 and 30/6/2020. Our analysis considered all variables regarding patients’ demographic characteristics and comorbidities that were systematically recorded in a national database. Discrepancies between studies from different countries and populations in data collection and reporting greatly limit the ability of reliable comparisons between findings. This is further reinforced by the intrinsic differences between population groups as well as by the different policies applied in each country with respect to the hospital admission and care of patients with COVID-19. In our study, we applied multivariable regression analysis to identify the risk factors associated with mortality among hospitalized patients with COVID-19. As this methodology can only adjust for measured confounders, there may be other unreported confounding factors that cannot be identified thus requiring further studies.

In conclusion, our novel finding is that allergies and previous influenza vaccination decrease mortality in COVID-19 patients. Further studies elucidating the effect of existing allergies and vaccination status may shed light in the immune responses associated with COVID-19 infection and assist in the identification of the underlying mechanisms. Most importantly, the verification of the beneficial effect of vaccination against influenza or other human viruses by more large scale epidemiological studies may significantly assist in the implementation of efficient protective measures against severe COVID-19 infection.

## Data Availability

Data will be made available on request

## What is already known on this subject?

Significant variations in COVID-19 morbidity and mortality rates have been recorded worldwide. Mortality from COVID-19 has been associated with older age, and the presence of comorbidities, primarily hypertension, diabetes, obesity, cardiovascular and respiratory disease in several populations.

## What this study adds?

Greece performed outstandingly well during the first wave of COVID-19 pandemic, minimizing the transmission of infection in the community through the timely application of strict public health measures that resulted in relatively small numbers of cases and related deaths. This is the first study to report mortality rates and predictive factors in hospitalized patients in the largest region of Attica. We found a beneficial effect of allergies and previous influenza vaccination against COVID-19 mortality, with potentially important implications in the understanding of disease pathophysiology and progression as well as in the implementation of preventive strategies.

## Notes

### Competing Interest Statement

The authors have declared no competing interest.

### Clinical Trial

Data were provided by the 1st Regional Health Authority of Attica, following the approval of the protocol by the Scientific Committee of the Greek Ministry of Health (27628 / 23-06-2020), ensuring legality of conduct, compliance with medical ethical standards and scientific validity.

### Funding Statement

No funding

